# Nature and dimensions of the cytokine storm and its attenuation by convalescent plasma in severe COVID-19

**DOI:** 10.1101/2020.09.21.20199109

**Authors:** Purbita Bandopadhyay, Ranit D’Rozario, Abhishake Lahiri, Jafar Sarif, Yogiraj Ray, Shekhar Ranjan Paul, Rammohan Roy, Rajshekhar Maiti, Kausik Chaudhuri, Saugata Bagchi, Ayan Maiti, Md. Masoom Perwez, Biswanath Sharma Sarkar, Devlina Roy, Rahul Chakraborty, Janani Srinivasa Vasudevan, Sachin Sharma, Durba Biswas, Chikam Maiti, Bibhuti Saha, Prasun Bhattacharya, Rajesh Pandey, Shilpak Chatterjee, Sandip Paul, Dipyaman Ganguly

**Author notes:** These authors made equal contribution to the study (Purbita Bandopadhyay, Ranit D’Rozario, Abhishake Lahiri, Jafar Sarif, Yogiraj Ray, Shekhar Ranjan Paul).

## Abstract

To characterize key components and dynamics of the cytokine storm associated with severe COVID-19 disease, we assessed abundance and correlative expression of a panel of forty eight cytokines in patients suffering from acute respiratory distress syndrome (ARDS), as compared to patients with mild disease. Then in a randomized control trial on convalescent plasma therapy (CPT) in COVID-19 ARDS, we analyzed the immediate effects of CPT on the dynamics of the cytokine storm as a correlate for the level of hypoxia experienced by the patients. Plasma level of monocyte chemotactic protein 3 was found to be a key correlate for clinical improvement, irrespective of therapy received. We also identified a hitherto unappreciated anti-inflammatory role of CPT independent of its neutralizing antibody content. Neutralizing antibodies as well as reductions in circulating interleukin-6 and interferon gamma induced protein 10, both contributed to marked immediate reductions in hypoxia in severe COVID-19 patients receiving CPT.

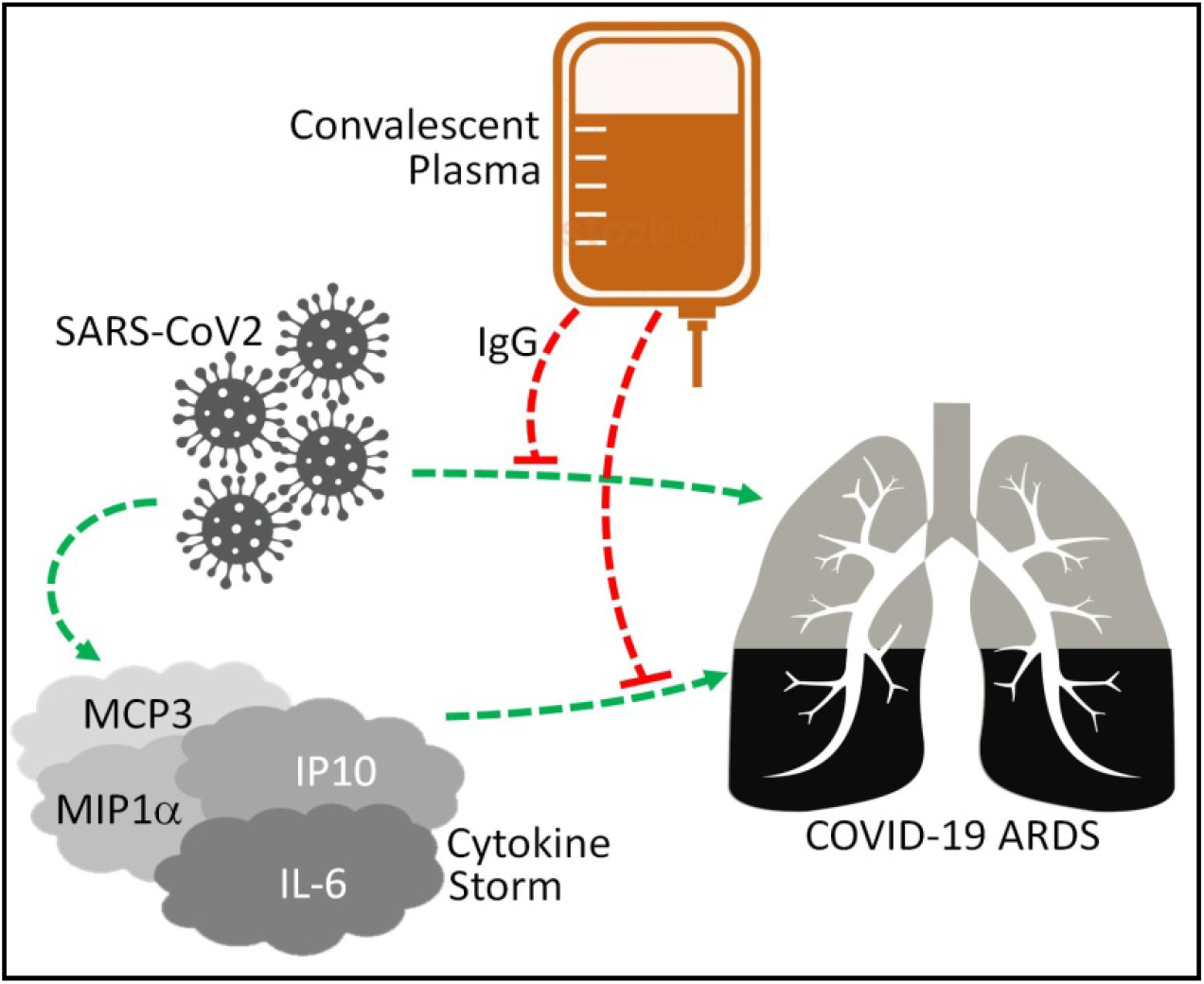

---

**T**he pandemic caused by the novel coronavirus SARS-CoV-2 has caused over 35 million infections and over 1 million deaths. The disease caused by SARS-CoV2, the coronavirus disease 2019 or Covid-19, have a spectrum of symptoms spread over two distinct phases in the symptomatic individuals. The symptomatology in the first milder phase variably includes malaise, fever, tussis, fatigue, cephalgia, myalgia, anosmia or ageusia followed by recovery (Huang et al., 2020). In a fraction of the infected individuals this milder phase progresses to a more severe disease mostly characterized by gradually worsening hypoxia requiring O2 supplementation and in some to acute respiratory distress syndrome (ARDS), leading to untoward fatal outcomes in a number of them (Huang et al., 2020; WHO, 2020). A hyper-immune activation response is associated with the severe symptoms, characterized by a systemic deluge of inflammatory cytokines or ‘cytokine storm’ (Laing et al., 2020; Arunachalam et al., 2020; Lucas et al., 2020). Different therapeutic approaches are currently being explored, either by repurposing specific anti-viral agents, viz. remdesivir (Goldman et al., 2020), or by using corticosteroids to affect immunomodulation (Horby et al., 2020). In addition, convalescent plasma therapy (CPT) has emerged as a widely tried strategy against COVID-19, being explored in a number of clinical trials all over the world (Joyner et al., 2020; Rubin, 2020; Li et al., 2020). Convalescent plasma transfusion is an age-old strategy for passive immunization, with the primary intention to supplement non-recovering patients with antibodies against specific pathogens (Rubin, 2020). First we aimed at an in-depth characterization of the nature and dimensions of the cytokine storm encountered in the COVID-19 patients progressing to ARDS, as compared to mild disease, and then, in a randomized control clinical trial on COVID-19 CPT, explored effect of convalescent plasma, if any, on mitigation of this systemic hyperimmune activation phenotype.

We recruited patients suffering from COVID-19 at the ID & BG Hospital, Kolkata, India, who either had mild COVID-19 disease (WHO Clinical Progression Score 1-4; N=13, 12 males and 1 female, aged 41.1 ± 11.2 years, enrolled after 2.5 ± 2 days after hospitalization) or more severe disease showing evidence for ARDS with PaO2/FiO2 ratio between 100-300 (WHO Clinical Progression Score 5-6; N=33, 26 males and 7 females, aged 60.03 ± 11.5 years, enrolled after 4 ± 2.8 days after hospitalization). As per our trial protocol, approved by Central Drugs Standard Control Organisation (CDSCO) India and registered with Clinical Trial Registry of India (No. CTRI/2020/05/025209), we randomized the recruited ARDS patients into either standard-of-care (SOC) group or added two doses of 200ml convalescent plasma (CP) on two consecutive days to their standard care (CPT). The first transfusion of ABO-matched convalescent plasma was done on the day of enrolment (day 1), followed by another transfusion on the next day (day 2). Plasma samples were taken on day 1 before CP transfusion (time-point 1 or T1) and again on day 3 or 4 post-transfusion (time-point 2 or T2) to assess the immediate effects of CP transfusion on the systemic cytokine milieu. All experiments were done with appropriate human ethical approvals from all involved institutions and after obtaining informed consent from the patients recruited.

First, we analyzed a panel of 48 cytokines in plasma at T1 for deeper characterization of the cytokine storm through comparison between patients having milder disease and patients with evidence of ARDS. We identified a panel of 14 molecules that were significantly higher in ARDS patients (Figure 1A, Supplemental figure 1). In addition, we found a significant decrease in plasma abundance of TNF-related apoptosis-inducing ligand (TRAIL) in ARDS, a cytokine known to be expressed in cytotoxic T cells and NK cells and involved in killing of virus-infected host cells (Waggoner et al., 2016), which may signify obviation of infected host cell-directed cytotoxicity at this later phase of the disease. Indeed, the viral load (estimated by average cycle threshold values from real time PCR for two viral genes from nasopharyngeal swabs, collected concomitant to plasma sampling) was significantly higher (lower cycle threshold values) in patients with milder disease, perhaps due to earlier sampling in their disease course (data not shown).

**Figure 1.**
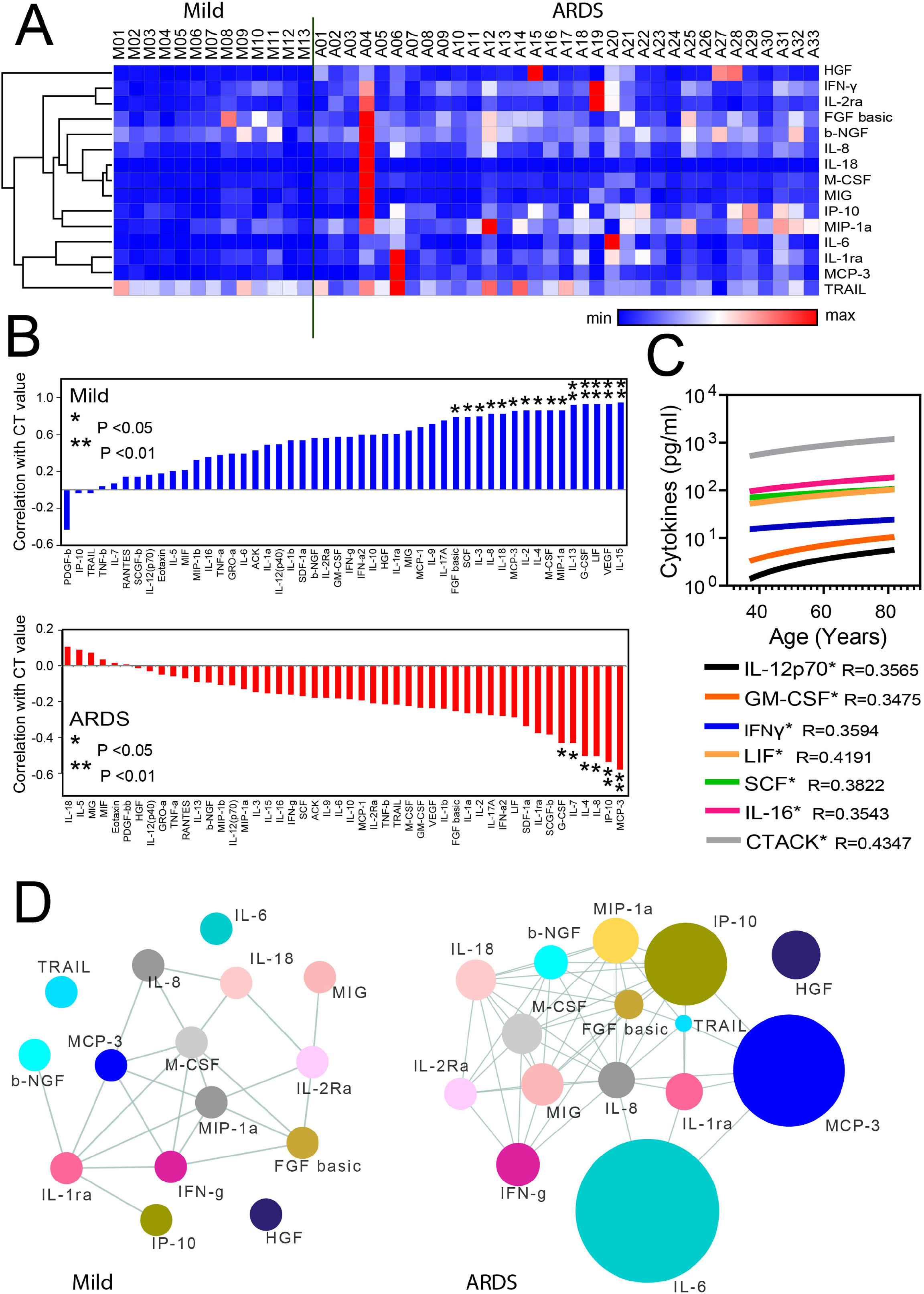
Nature and dimensions of the cytokine storm in severe Covid-19 disease. **A**. Heatmap representing normalized plasma abundance of 15 majorcytokines compared between patients with mild disease (n=13) or ARDS (n=33) at enrolment, made with Morpheus. Rows are clustered using one minus Pearson correlation distance and average linkage. **B**. Extent of correlation between the plasma levels of cytokines and concomitant viral loads as measured by average cycle threshold values for real time PCR of two SARS-CoV2 target genes, compared between patients with mild disease and ARDS. Spearman correlation values are arranged in ascending order and significance are marked by either * (P<0.05) or ** (P<0.01). **C**. Plasma level of cytokines having significant (P<0.05) correlation with age of patients with ARDS. The Spearman R value is given for each of the colour coded cytokine with age. **D**. Correlation network of major cytokines at T1 for both mild disease group (n=13) and ARDS group (n=33). Pearson correlations with R> 0.3 and P<0.05 are considered.

Plasma abundance of a number of cytokines in the mild disease was negatively correlated with concomitant viral load, presumably representing the cytokine component of an efficient protective immunity in the milder phase (Figure 1B). Interestingly, in patients with ARDS there was rather a significant positive correlation of viral load with a few specific cytokines (Figure 1B). Among them interleukin-8 (IL-8) and granulocyte colony stimulating factor (G-CSF) presumably represent the usual neutrophil recruitment response triggered by residual virus-infected cells. Notably most of the major cytokine storm components, viz. interleukin-6 (IL-6), interferon-γ (IFNγ), interleukin-1 receptor antagonist (IL-1RA), macrophage inflammatory protein 1α (MIP1α), seemed not to correlate any way with the concomitant viral load in patients with ARDS. But notable exception here were monocytes chemotactic protein 3 (MCP3) and interferon gamma induced protein 10 (IP10), which may represent pathogenic links between host-virus interactions in the earlier phase of the disease and the hyper-immune activation that ensue later in a fraction of patients and warrant further mechanistic studies. Of note here, increasing age of the patients was significantly correlated with higher plasma abundance of a number of cytokines, though except IFNγ none of them were significantly associated with ARDS (Figure 1C).

Next to gather some insight on the dimensions of the cytokine storm encountered in COVID-19 ARDS, we constructed the correlative networks (Pearson correlation, with a cut-off of R >0.5, p<0.01) among the individual members of the whole panel of cytokines and compared between mild and ARDS patients (Supplemental figure 2A,B). We also separately constructed similar correlative networks (Pearson correlation, with a cut-off of R >0.5, p<0.05) among the cytokines that were found to be significantly dysregulated in ARDS (Figure 1D). Interestingly, in ARDS we found more robust assimilation of correlative networks among the cytokines with greater number of edges compared to mild disease (156 edges in mild vs 356 edges in ARDS among the whole panel of 48 cytokines and 23 edges in mild disease vs 52 edges in ARDS among the cytokines significantly dysregulated in ARDS). In this analysis the most notable was a five member cytokine module, comprising of IL-6, monocytes chemotactic protein 3 (MCP3), MIP1α, IL-1RA and interferon gamma induced protein 10 (IP10), showing robust correlative upregulation. On the other hand, a notable omission from this correlative network, despite a significantly higher abundance in ARDS, was that of the mesenchymal cell-derived pleiotropic cytokine hepatocyte growth factor (HGF), perhaps representing an anti-inflammatory as well as tissue regeneration response at the face of the systemic inflammatory assault, as shown earlier (Lee et al., 2020). Although a proinflammatory role of HGF, targeting neutrophils through c-MET receptor signaling, has also been described recently and thus may be of interest to explore in the context of severe COVID-19 (Stakenborg et al., 2020).

We then analyzed the aforementioned cytokine panel in all ARDS patients at T2 and explored if the T1 to T2 change was different between SOC and CPT groups. Intriguingly, on analyzing the panel of 15 cytokines depicted in Figure 1A, we found a notable effect of CPT, as compared to SOC, in reducing the levels of a number of them, viz. IL-6, IP10 and macrophage colony stimulating factor (MCSF) (Figure 2A,B; Supplemental table 1). On the other hand, none of the major cytokines driving the cytokine storm was found to be significantly reduced at T2 in patients receiving SOC (Supplemental table 1). This anti-inflammatory effect was not dependent on either the anti-SARS-CoV-2 spike Immunoglobulin G (IgG) content of the transfused convalescent plasma or its capacity for blocking the interaction between SARS-CoV2 spike protein and angiotensin converting enzyme 2 (ACE2) by CP (Figure 2A). Of note here, we found strong correlation between anti-spike IgG content of plasma and its capacity to neutralize spike-ACE2 interaction, as measured in an in vitro assay (Tan et al., 2020), in our cohort of convalescent donors (R=0.8564, P<0.0001).

**Figure 2.**
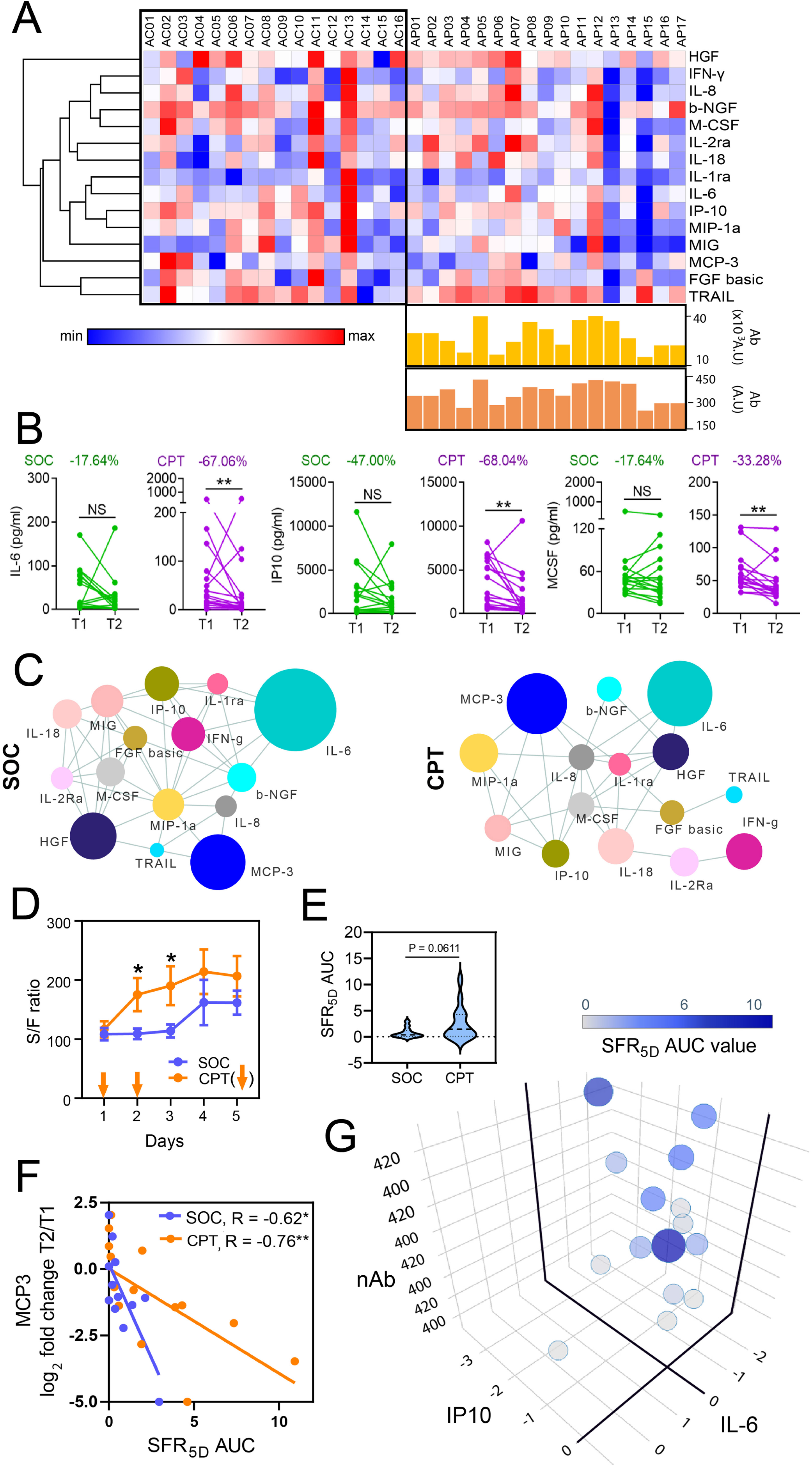
Attenuation of cytokine storm and mitigation of hypoxia in response to convalescent plasma. **A**. Heatmap representing log2 fold change between T1 and T2 in the plasma abundance of major cytokines associated with ARDS compared between SOC (n=16) and CPT (n=17) groups. Heatmap is made with Morpheus and rows are clustered using one minus Pearson correlation distance and average linkage. Anti-Spike IgG content and nAb content of the convalescent plasma transfused are represented for the respective patients in the CPT group. **B**. Reduction in plasma levels of IL-6, IP10 and M-CSF from T1 to T2, compared between SOC and CPT groups. The percentage difference of median value at T2 time point in compare to T1 was also calculated for three of the cytokines. **C**. Correlation network of major cytokines at T2 compared between SOC (n=16) and CPT (n=17) groups. Pearson correlations with R> 0.3 and P<0.05 are considered. **D**. Kinetics of SpO2/FiO2 ratio in patients over five days post-enrolment, compared between SOC and CPT groups. Comparison of day means was performed by Welch’s t-test and significant differences are marked by * (P<0.05). The plasma transfusion days are also marked by arrows. **E**. Violin plot represents the comparison of SFR_5D_AUC for the patients between SOC and CPT groups. **F**. Correlation of log2 fold change between T1 and T2 for plasma level of MCP3 and SFR_5D_AUC. The Spearman correlations values for SOC and CPT groups are given with their significance level. **G**. SFR_5D_AUC relationships with nAb content of transfused plasma, log2 fold change of plasma levels of IL-6 and IP10 in the CPT group. The plot is generated by plotly package in R.

This anti-inflammatory effect of convalescent plasma was also apparent when we compared the correlative networks at T2 of both the whole panel of cytokines (Supplemental figure 3 A,B) as well as among the 15 cytokines found to be significantly dysregulated in ARDS (Fig. 2C) and compared them between the SOC and CPT groups. The residual correlative edges among the cytokines at T2 were significantly less in number in case of CPT in contrast to SOC group (334 edges in SOC group vs 135 edges in CPT group among the whole panel of 48 cytokines and 47 edges in SOC group vs 33 edges in CPT group among the cytokines significantly dysregulated in ARDS), representing an attenuation of the cytokine storm in response to CPT and a trend toward a cytokine milieu quite similar to one found in the early milder phase of the disease.

To explore if the effect of therapy on the cytokine storm was correlated with concomitant clinical status of the ARDS patients, we decided to assess their requirement for oxygen supplementation (as measured by fraction of inspired oxygen or FiO2) for maintaining O2 saturation of circulating hemoglobin at a physiological level (O2 saturation as measured by pulse oximetry or SpO2), as this offered a quantifiable, comparable and relevant clinical parameter across all ARDS patients. Thus the immediate clinical outcome in the ARDS patients were assessed by the kinetics of SpO2/FiO2 ratio (S/F ratio) for 5 days following enrolment (Figure 2D). This data was then processed to represent clinical improvement over 5 days with respect to day 1 by calculating the area under curve (AUC) for the S/F Ratio curve (SFR_5D_AUC) (Figure 2E). CPT was found to affect faster mitigation of hypoxia, as compared to patients receiving SOC only (Figures 2D,E). We noted gradual abrogation of this differential response on reductions in hypoxia third day onwards after 2^nd^ dose of CPT, indicating that a sustained benefit may require additional transfusions of CPT in some patients.

Reduction in the chemokine MCP3 at T2, as compared to T1 was significantly associated with the mitigation of hypoxia, irrespective of whether patients received CPT or not, thus identifying a major pathogenic molecule underlying COVID-19 ARDS (Figure 2F). In patients receiving CPT, as expected, the improvements in S/F ratio was significantly correlated with neutralizing antibody (nAb) content of the convalescent plasma they were transfused with. Interestingly, in a linear regression analysis we identified that IL-6 and IP10, the two major ARDS-associated cytokines that were affected by the anti-inflammatory effect of CPT as described in Figure 2A, also played a major role in this immediate mitigation of hypoxia in combination with nAb content (Figure 2G; Supplemental table 2). Thus beneficial effect of CPT perhaps mechanistically goes beyond just passive immunization of the recipients and thus should further be explored mechanistically to identify other anti-inflammatory factors in convalescent plasma. We envisage that this anti-inflammatory effect of CPT may affect mitigation of other longer term systemic effects of the cytokine storm encountered in severe COVID-19, full appreciation of which awaits end-point analyses in our trial as well as further meta-analyses of data from other clinical trials on CPT accomplished or ongoing elsewhere.

Thus, this study characterizes the nature and dimensions of the hyper-immune activation-associated cytokine storm in patients suffering from acute respiratory distress syndrome, which could identify a number of hitherto unappreciated features of the disease pathogenesis in severe COVID-19. Moreover, we report here an anti-inflammatory effect of COVID-19 convalescent plasma, independent of, but acting in synergy with, its neutralizing antibody content, which may prove to be a composite predictor of response to convalescent plasma therapy in COVID-19 and should be explored while analyzing the clinical outcomes of trials ongoing throughout the world.

## Data Availability

All data are available with the corresponding author and will be shared on request.

## Acknowledgements

D.G. acknowledges funding for the RCT and associated immune monitoring studies from Council of Scientitific Industrial Research (CSIR), Govt. of India (MLP-129); R.P. acknowledge funding from CSIR (MLP-2005) and Fondation Botnar. D.G. holds a Swarnajayanti Fellowship from Department of Science & Technology (DST), Govt. of India, S.P. holds a Ramanujan Fellowship from Science & Engineering Research Board (SERB), Govt. of India, S.C. holds an Intermediate Fellowship from DBT-Welcome Trust India Alliance and R.P. holds a Ramalingaswami Fellowship from Department of Biotechnology (DBT), Govt. of India. P.B. and A.L. are supported by Senior Research Fellowship from CSIR, R.D. and J.S. are supported by Junior Research Fellowships from University Grants Commission, India. Authors express their gratitude to Anurag Agrawal and Kamakshi Sureka for critical reading of the manuscript and Shantanu Sengupta for help with neutralizing antibody assay.

## Author contributions

D.G. conceptualized the study. D.G., S.P. and Y.R. designed the study protocol. P.Ba. and R.D. did the plasma cytokine analysis. J.S. performed serological studies. A.L. contributed to the analysis and computation. Y.R. and S.R.P. recruited patients, maintained clinical data and supervised clinical management. R.R., R.M., K.C., S.B., A.M., M.M.R., B.S.S., D.R., R.C., B.S. contributed to patient management. J.S.V., S.S. and R.P. did the RT-PCR for SARS-CoV2. S.C. contributed in immunological studies. S.P. and P.Bh. recruited convalescent donors, D.B., C.M. and P.Bh. performed donor screening, apheresis and biobanking of convalescent plasma. D.G. and S.P. wrote the manuscript. All authors approved the manuscript.

## Supplemental methods

### Ethical approval

The randomized control trial (RCT) on convalescent plasma therapy and all associated studies and the human sampling needed for them were done with informed consent from the patients as per recommendation and ethical approval from the Institutional Review Boards of CSIR-Indian Institute of Chemical Biology, Kolkata, India (IICB/IRB/2020/3P), Institutional Review Boards of Medical College Hospital, Kolkata (MC/KOL/IEC/NON-SPON/710/04/2020), India and Infectious Disease & Beleghata General Hospital (ID & BG Hospital), Kolkata, India (IDBGH/Ethics/2429). The RCT was approved by Central Drugs Standard Control Organisation (CDSCO) under Directorate General of Health Services, Ministry of Health & Family Welfare, Govt. of India (approval no. CT/BP/09/2020) and registered with Clinical Trial Registry of India (CTRI), under Indian Council of Medical Research, India.

### Plasma cytokine analysis

Plasma was isolated from peripheral blood of patients collected in EDTA vials and cytokine levels (pg/ml) were measured using the Bio-Plex Pro Human Cytokine Screening Panel 48-Plex Assay (Bio-Rad, Cat No. 12007283) which quantitates 48 cytokines (FGF basic, Eotaxin, G-CSF, GM-CSF, IFN-γ, IL-1β, IL-1ra, IL-1α, IL-2Rα, IL-3, IL-12 (p40), IL-16, IL-2, IL-4, IL-5, IL-6, IL-7, IL-8, IL-9, GRO-α, HGF, IFN-α2, LIF, MCP-3, IL-10, IL-12 (p70), IL-13, IL-15, IL-17A, IP-10, MCP-1 (MCAF), MIG, β-NGF, SCF, SCGF-β, SDF-1α, MIP-1α, MIP-1β, PDGF-BB, RANTES, TNF-α, VEGF, CTACK, MIF, TRAIL, IL-18, M-CSF and TNF-β). The plasma samples were diluted (1:3) in sample diluent and the assay was performed using manufacturer’s protocol. The plate was run and analyzed using Bio-Plex® 200 System (Bio-Rad).

### RNA Isolation from COVID-19 Samples in TRIzol

RNA from COVID-19 samples in TRIzol samples were extracted using chloroform-isopropanol method. 1/5th volume of chloroform (Cat No. 1024452500, Merck) was added to the TRIzol (Cat No. 15596026, Invitrogen) and mixed thoroughly followed by 5 minutes incubation at room temperature. After centrifugation at 12,000 rpm at 4°C for 15 minutes, aqueous phase was transferred to a fresh microfuge tube. To precipitate RNA, 2-Propanol (Cat No. 1096341000, Merck) was added in equal ratio and incubated on ice for 10 minutes followed by centrifugation at 10,000 rpm at 4°C. RNA pellet was washed twice with 75% ethanol (Cat No. 100983, Merck). The pellet was re-suspended in 20 µL of RNase free water (Cat No. AM9920, Thermo Fisher Scientific) followed by quantitation using NanoDrop (Cat No. ND-2000, Thermo Fisher Scientific).

### RT-PCR

qRT-PCR for SARS-CoV-2 detection was performed using the STANDARD M nCoV Real-Time Detection kit (Cat No. 11NCO10, SD Biosensor), approved by Indian Council of Medical Research (ICMR), India. Briefly, 5 µL of RNA was added to the reaction mix, as per manufacturer’s protocol. The RT-PCR was run on QuantStudio 6 Flex Real-Time PCR Systems (Applied Biosystems, Thermo Fisher Scientific) using recommended cycling conditions in a 96 well format. A cycle threshold (Ct-values) cut-offs mean value for both RdRp and E gene was considered as per SD biosensor’s manual for interpreting the results. A CY5 labeled Internal Control was used.. Average CT values for two SARS-CoV2 targets (RdRp and E) were used for the study as a surrogate for viral load.

### SARS-CoV-2 Surrogate Virus Neutralization Assay

Neutralizing antibodies against SARS-CoV-2 in human plasma samples from peripheral blood of convalescent donors were detected using GeneScript SARS-CoV-2 Surrogate Virus Neutralization kit (Cat no-L00847). The kit consists of recombinant SARS-CoV-2 Spike protein receptor binding domain fragment conjugated with horseradish peroxidase (HRP-RBD) & human ACE2 receptor protein (hACE2). Presence of SARS-CoV-2 neutralizing antibodies in the plasma samples block interaction between HRP-RBD and hACE2, which is detected through colorimetry. Assay was performed according to manufacturer’s protocol. Plasma samples, and provided positive and negative controls were diluted at a ratio of 1:10 with the sample dilution buffer. HRP-RBD solution was added to the diluted plasma samples and controls at a ratio of 1:1 (vol/vol) and incubated at 37°C for 30 minutes. The 100µl of the above diluted samples and controls were added to the wells and further incubated at 37°C for 15 minutes before wash and substrate addition. Absorbance value was recorded following addition of stop solution at 450nm using Bio-RAD iMARK microplate reader. Since presence of SARS-CoV-2 neutralizing antibodies in the plasma samples will result in inhibition of interaction between HRP-RBD and plate-bound human ACE2 protein, and subsequent development of color, assay results are interpreted as inhibition rate of assay reaction. Inhibition rate is calculated as follows: Inhibition = {1-(O.D value of sample/O.D value of negative control)} × 100 [Inhibition values ≥20% signify positive detection of neutralizing antibodies]. In case of recipient patients the amount of neutralizing antibodies was calculated considering the amount (ml) of plasma transfused to them.

### ELISA for anti SARS-CoV-2 IgG

Levels of Immunoglobulin G (IgG) specific for SARS-CoV-2 in the plasma isolated from peripheral blood of recovered patients were detected using EUROIMMUN Anti-SARS-CoV-2 (IgG) Elisa kit (Cat No-EI 2606-9601 G). This assay provides semiquantitative estimation of IgG levels against SARS-CoV-2 spike protein. Assay was performed according to manufacturer’s protocol. The assay wells are pre-coated with recombinant S1 domain of the SARS-CoV-2 spike protein. Plasma samples were diluted with provided sample dilution buffer at a ratio of 1:101 (vol/vol). The two reaction steps in this ELISA involves the incubation of diluted patient samples in pre-coated wells following which bound antibodies are detected using enzyme labeled anti human IgG. O.D. was measured at 450nm and reference wavelength of 655nmusing Bio-RAD iMARK microplate reader. Presence of anti SARS-CoV-2 IgG antibodies in the plasma was measured using the following formula: Ratio = Extinction of the control or patient samples/Extinction of calibrator (Ratio ≥ 1.1 is interpreted as positive). In case of recipient patients the amount of IgG was calculated considering the amount (ml) of plasma transfused to them.

### Standard-of-care

All patients infected with SARS-CoV2 at diagnosis either at ER or OPD or through Telemedicine while being in home isolation had received either of these two combinations: Hydroxychloroquine 400 mg BD on first day followed by 400 mg OD for four days plus Azithromycin 500mg OD for 5 days or Ivermectin 12 mg OD for 5days plus Doxycyclin 100 mg BD for 10 days. At the clinical trial site (ID & BG Hospital, Kolkata, India) standard-of-care (SOC) in all patients with evidence for ARDS were: O2 therapy as per requirement in all patients, dexamethasone or equivalent corticosteroid in all patients, for patients with D-dimer <1000 Fibrinogen Equivalent Units (FEU) prophylactic anticoagulation and for patients with D-dimer >1000 ng/ml FEU therapeutic anticoagulation using either low molecular weight heparin or unfractionated heparin, appropriate broad-spectrum antibiotic therapy based on lymphocyte count, clinical and microbiological assessment, blood sugar was maintained below 200mg/dl using appropriate anti-diabetic therapy, appropriate anti-hypertensive agents were used as per requirement to maintain systolic blood pressure 100-140 mm of Hg, diastolic blood pressure at 70-90 mm of Hg and mean arterial pressure >65 mm of Hg. Awake proning for 6-8 hours/day was attempted in all patients with ARDS. O2 therapy was designed to maintain SpO2 >95% using different devices with different efficiencies in O2 supplementation (as denoted by FiO2), viz. nasal canula, face mask, face mask with reservoir, and in patients unable to maintain SpO2 above 90% with face mask with reservoir, high flow nasal cannula (HFNC) or in some cases mechanical ventilation (MV). For S/F ratio kinetics, a value of 89.99 was used for data-points where either HFNC or MV was in use. Of note S/F ratio kinetics was analyzed only in patients having data for at least one intervening time-point in addition to day 1 and day 5.

### Convalescent plasma therapy

Plasma was collected from convalescent donors (recovered from RT-PCR positive SARS-CoV-2 infection at least 28 days prior to donation) by apheresis at the Department of Blood Transfusion and Immunohematology, Medical College Hospital, Kolkata, India. All donors were tested for their anti-Spike IgG content in addition to routine screening tests to exclude major blood borne pathogens before apheresis. As per our trial protocol (Clinical Trial Registry of India No. CTRI/2020/05/025209), approved by Central Drugs Standard Control Organisation (CDSCO) India, we randomized the ARDS patients into either standard-of-care (SOC) group as controls or added two consecutive doses of ABO-matched 200ml convalescent plasma on two consecutive days to their standard care (CPT group), the first transfusion being on the day of recruitment (Day 1).

### Co-occurrence analyses

Co-occurrence among each pair of cytokines was calculated using Pearson correlation (r) and corresponding p-value of the correlation was measured using a t-distribution. Absolute values of the cytokines were used for the calculation of correlation network and threshold was set to r>0.5, p<0.01 for the complete set of cytokines from mild (n=13) and ARDS (n=33) conditions while the threshold was set to r>0.3, p<0.05 for set of significantly different cytokines between mild and ARDS conditions in SOC (n=16) and Plasma (n=17) sets at T2. All calculations were done using ‘Hmisc’ R package and finally converted to a network file using the ‘igraph’ R package. Visualisation of the network was performed using Cytoscape. Each cytokine was color coded and node size was set in proportion to the median fold change as compared to the same cytokine in the mild datasets.

### Statistical analyses

Differential abundance of cytokines was evaluated by Mann-Whitney U Test and Wilcoxon Matched Pairs Test for categorical and ordinal data, respectively. In order to test the monotonous relationship between CT value and cytokine abundance of the patients, spearman’s rank correlation was used, followed by p-value calculation using t-distribution. Similar tests were performed between neutralizing antibody content and IgG antibody content for plasma recipient patients. In order to explain the SFR_5D_ AUC as a function of a combination of cytokines and nAb, linear regression was applied on the ranks of each of the parameters and was implemented using R. The statistical significance was defined as a p value <0.05 (*) and p value <0.01 (**) with two-sided tests, unless otherwise mentioned. Statistical tests were also performed in Statistica 64 (StatSoft).

## Legends for supplemental figures and tables

**Supplemental Figure 1:**
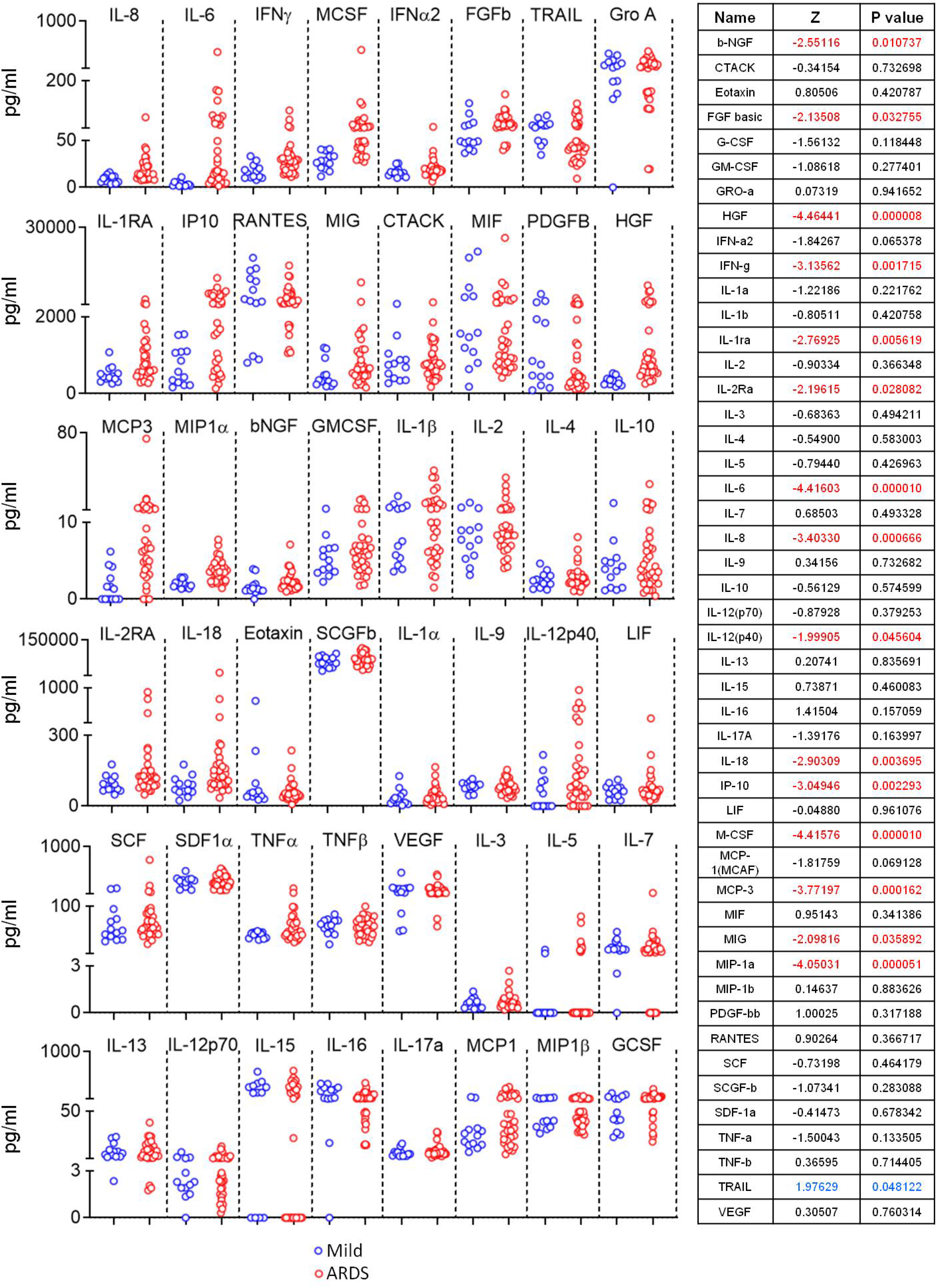
Differential levels of cytokines between mild and ARDS samples. Levels of 48 measured cytokines from 46 patient samples were compared between mild and ARDS groups. Blue and red dots represent mild and ARDS samples respectively. The Z value and P value for Mann-Whitney U Test between the groups are given in the adjacent table. Significantly higher and lower differences in cytokine levels for ARDS group are mentioned by red and blue fonts respectively.

**Supplemental Figure 2:**
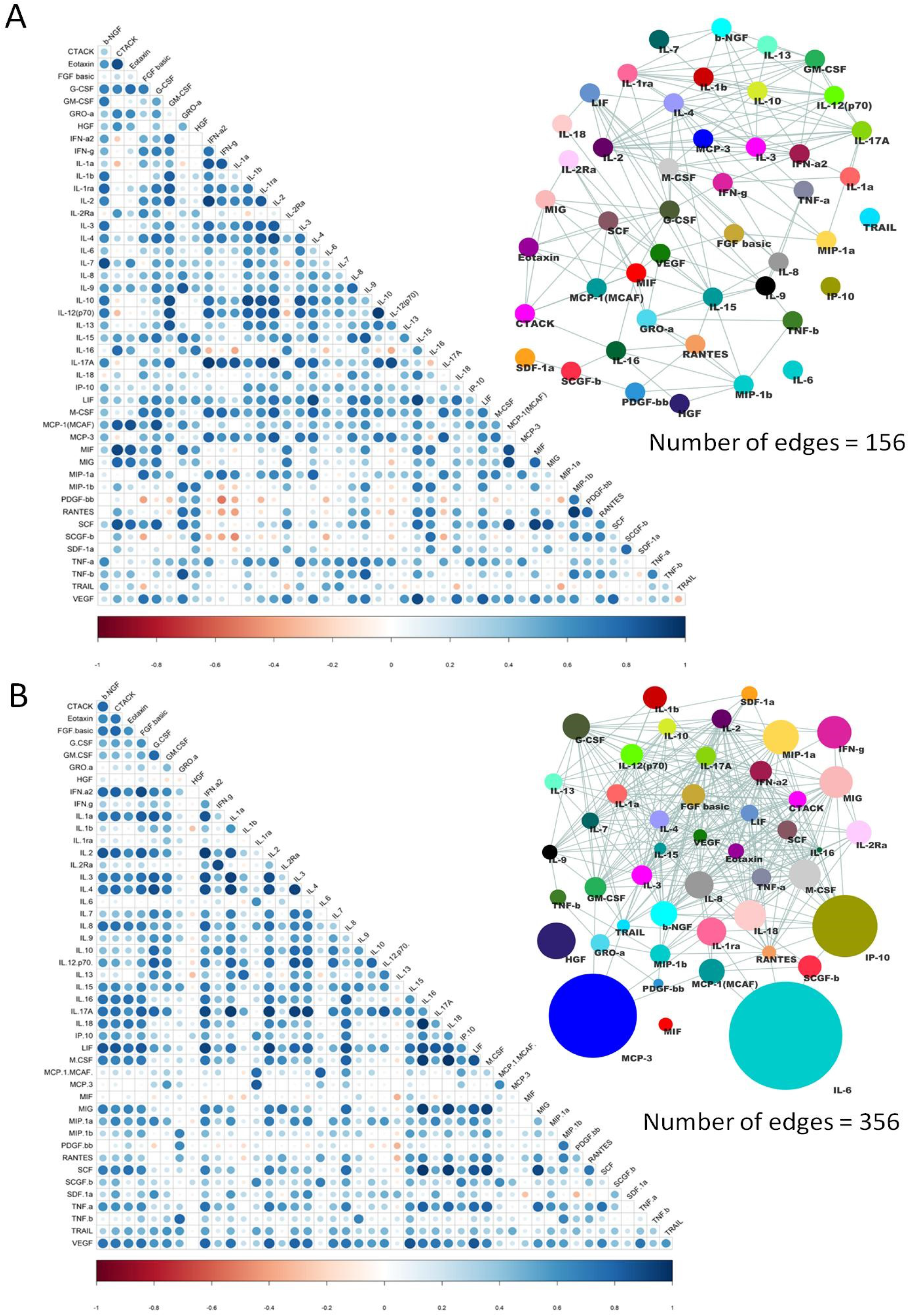
Correlations of cytokine levels across the samples for at T1 time point for mild and ARDS groups and at T2 time point for SOC and CPT groups. 2(A) and (B) describe the correlation network at T1 time point in mild group (n=13) and in ARDS group (n=33) respectively. Whereas, 3(A) and (B) represent the correlation network at T2 time point in SOC group (n=16) and in ARDS group (n=17) respectively. Pearson correlation values were calculated for each comparison. The minimum correlation value was set at 0.5 and subsequent P value threshold was <0.01. All calculations were done using ‘Hmisc’ R package and finally converted to a network file using the ‘igraph’ R package. Visualisation of the network was performed using Cytoscape. Each cytokine was colour coded and node size was set in proportion to the median fold change as compared to the same cytokine in the mild datasets.

**Supplemental Figure 3:**
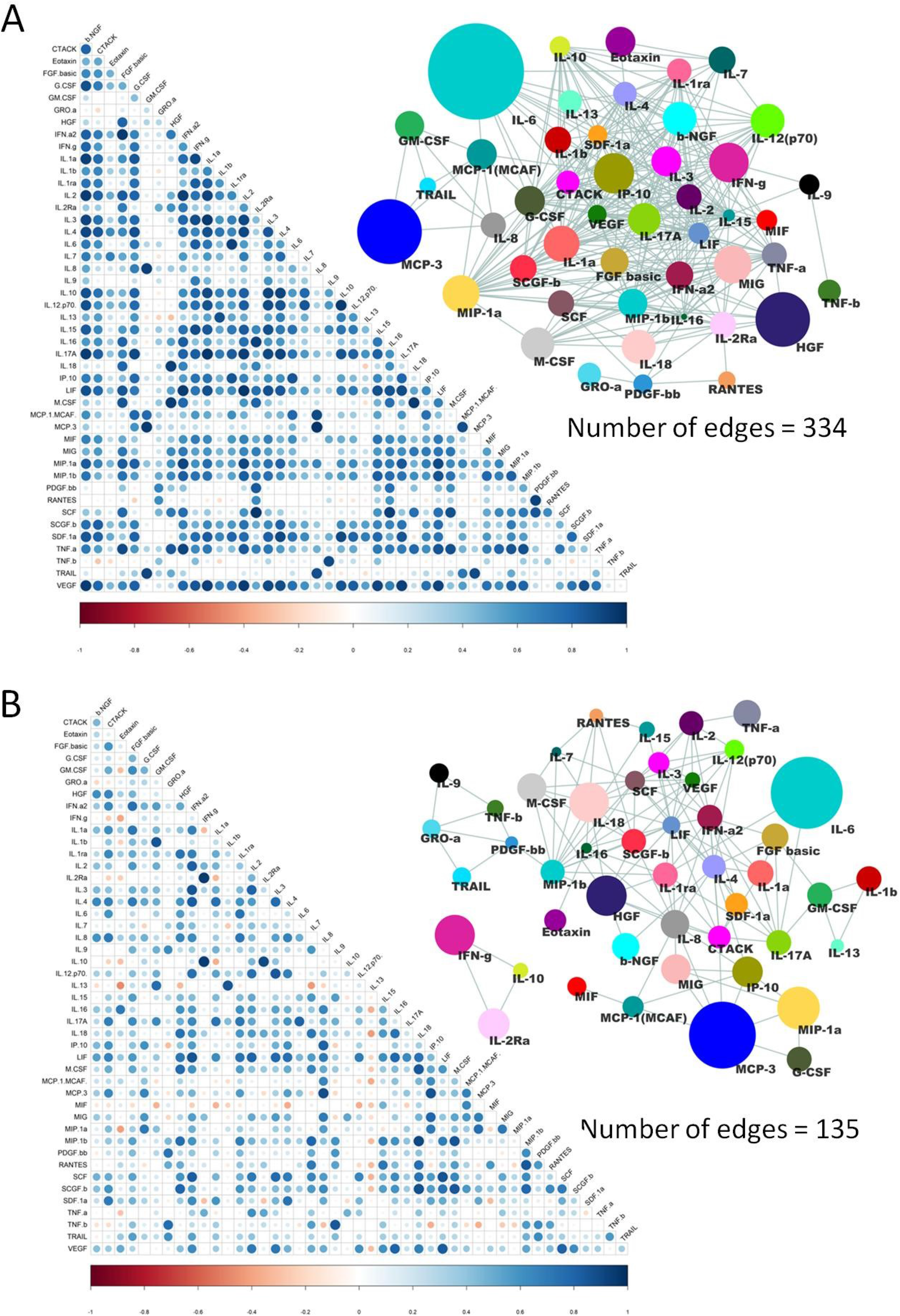
Correlations of cytokine levels across the samples for at T1 time point for mild and ARDS groups and at T2 time point for SOC and CPT groups. 2(A) and (B) describe the correlation network at T1 time point in mild group (n=13) and in ARDS group (n=33) respectively. Whereas, 3(A) and (B) represent the correlation network at T2 time point in SOC group (n=16) and in ARDS group (n=17) respectively. Pearson correlation values were calculated for each comparison. The minimum correlation value was set at 0.5 and subsequent P value threshold was <0.01. All calculations were done using ‘Hmisc’ R package and finally converted to a network file using the ‘igraph’ R package. Visualisation of the network was performed using Cytoscape. Each cytokine was colour coded and node size was set in proportion to the median fold change as compared to the same cytokine in the mild datasets.

**Supplemental table 1:** Variations in the levels of 15 significantly differential cytokines between mild and ARDS conditions in SOC and CPT patients from T1 to T2 time points. The percentage difference of median value at T2 time point in compare to T1 was calculated for each of the cytokines. * P < 0.05, ** P < 0.01.

| Cytokines | Change in Median value from T1 to T2 (%) |  |
| --- | --- | --- |
|  | SOC | CPT |
| b-NGF | -3.37 | -22.89 |
| FGF basic | -12.80 | -9.57 |
| HGF | -1.55 | -19.83 |
| IFN-g | -17.33 | -7.77 |
| IL1-ra | -26.53 | -39.24 |
| IL2-ra | -23.23 | -5.18 |
| IL6 | -28.67 | -67.06* |
| IL8 | -21.51 | -37.28 |
| IL18 | -17.65 | -10.47 |
| IP10 | -47.00 | -68.04** |
| M-CSF | -17.64 | -33.28** |
| MCP-3 | -42.62 | -42.15 |
| MIG | 1.48 | -44.79 |
| MIP-1a | -3.54 | -15.04 |
| TRAIL | -17.92 | 7.54 |

**Supplemental table 2:**
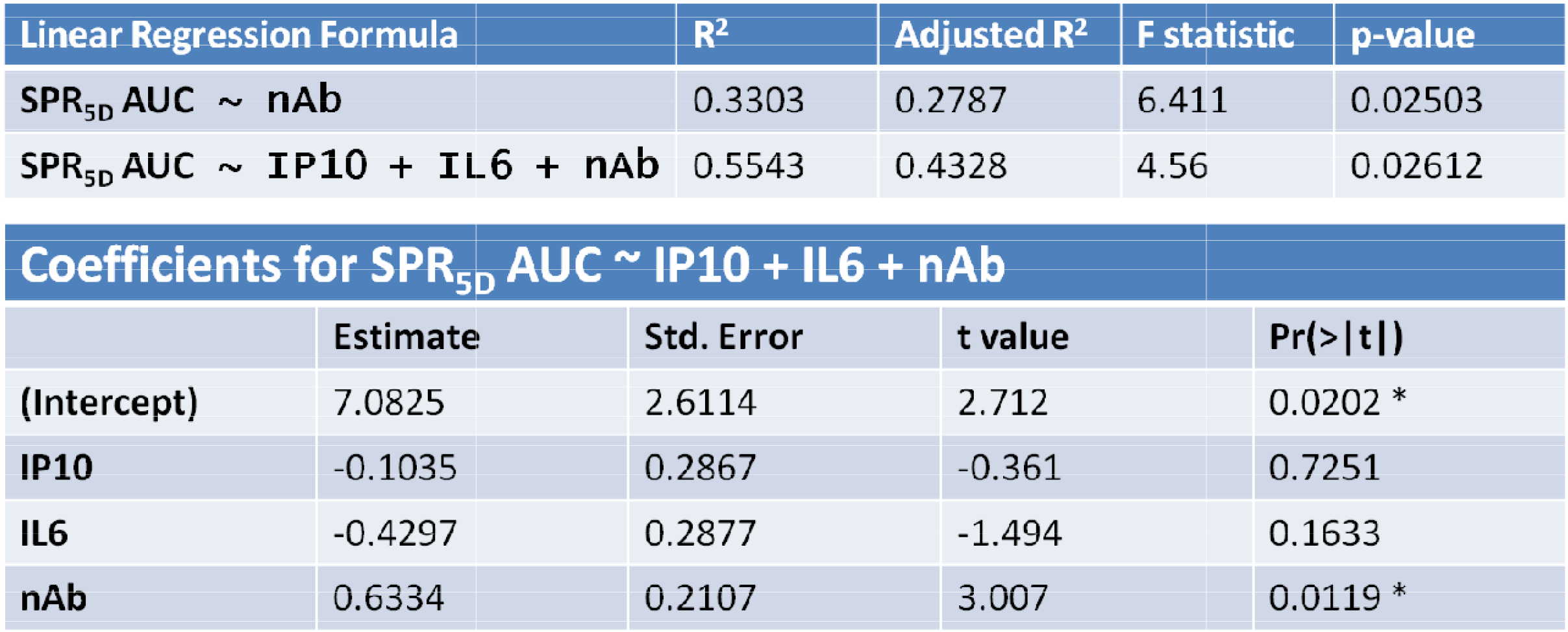
Linear regression analysis. Upper table represents the parameters for different linear regression models for SPR_5D_ AUC. The lower table represents the coefficients of multiple linear regression model for SPR_5D_ AUC on ranked values of IP10, IL6 and nAb. The analysis was performed using R.

## References

Arunachalam, P.S., Wimmers, F., Mok, C.K.P., Perera, R.A.P.M, Scott, M., Hagan, T., Sigal, N., Feng, Y., Bristow, L., Tak-Yin Tsang, O., Wagh, D., Coller, J., Pellegrini, K.L., Kazmin, D., Alaaeddine, G., Leung, W.S., Chan, J.M.C., Chik, T.S.H., Choi, C.Y.C., Huerta, C., Paine McCullough, M., Lv, H., Anderson, E., Edupuganti, S., Upadhyay, A.A., Bosinger, S.E., Maecker, H.T., Khatri, P., Rouphael, N., Peiris, M., Pulendran, B. (2020). Systems biological assessment of immunity to mild versus severe COVID-19 infection in humans. Science. 369, 1210–1220.

Goldman, J.D., Lye, D.C.B., Hui, D.S., Marks, K.M., Bruno, R., Montejano, R., Spinner, C.D., Galli, M., Ahn, M.Y., Nahass, R.G., Chen, Y.S., SenGupta, D., Hyland, R.H., Osinusi, A.O., Cao, H., Blair, C., Wei, X., Gaggar, A., Brainard, D.M., Towner, W.J., Muñoz, J., Mullane, K.M., Marty, F.M., Tashima, K.T., Diaz, G., Subramanian, A.; GS-US-540-5773 Investigators. (2020) Remdesivir for 5 or 10 Days in Patients with Severe Covid-19. N. Engl. J. Med. doi: 10.1056/NEJMoa2015301.

Horby, P., Lim, W.S., Emberson, J.R., Mafham, M., Bell, J.L., Linsell, L., Staplin, N., Brightling, C., Ustianowski, A., Elmahi, E., Prudon, B., Green, C., Felton, T., Chadwick, D., Rege, K., Fegan, C., Chappell L.C., Faust, S.N., Jaki, T., Jeffery, K., Montgomery, A., Rowan, K., Juszczak, E., Baillie, J.K., Haynes, R., Landray, M.J. (2020) Dexamethasone in Hospitalized Patients with Covid-19 - Preliminary Report. N. Engl. J. Med. doi: 10.1056/NEJMoa2021436.

Huang, C., Wang, Y., Li, X., Ren, L., Zhao, J., Hu, Y., Zhang, L., Fan, G., Xu, J., Gu, X., Cheng, Z., Yu, T., Xia, J., Wei, Y., Wu, W., Xie, X., Yin, W., Li, H., Liu, M., Xiao, Y., Gao, H., Guo, L., Xie, J., Wang, G., Jiang, R., Gao, Z., Jin, Q., Wang, J., Cao, B. (2020) Clinical features of patients infected with 2019 novel coronavirus in Wuhan, China. Lancet. 395, 497–506.

Joyner, M.J., Senefeld, J.W., Klassen, S.A., Mills, J.R., Johnson, P.W., Theel, E.S., Wiggins, C.C., Bruno, K.A., Klompas, A.M., Lesser, E.R., Kunze, K.L., Sexton, M.A., Diaz Soto, J.C., Baker, S.E., Shepherd, J.R.A., van Helmond, N., van Buskirk, C.M., Winters, J.L., Stubbs, J.R., Rea, R.F., Hodge, D.O., Herasevich, V., Whelan, E.R., Clayburn, A.J., Larson, K.F., Ripoll, J.G., Andersen, K.J., Buras, M.R., Vogt, M.N.P., Dennis, J.J., Regimbal, R.J., Bauer, P.R., Blair, J.E., Paneth, N.S., Fairweather, D., Wright, R.S., Carter, R.E., Casadevall, A. (2020) Effect of Convalescent Plasma on Mortality among Hospitalized Patients with COVID-19: Initial Three-Month Experience. medRxiv. doi: 10.1101/2020.08.12.20169359

Laing, A.G., Lorenc, A., Del Barrio, I., Das, A., Fish, M., Monin, L., Muñoz-Ruiz, M., McKenzie, D.R., Hayday, T.S., Francos-Quijorna, I., Kamdar, S., Joseph, M., Davies, D., Davis, R., Jennings, A., Zlatareva, I., Vantourout, P., Wu, Y., Sofra, V., Cano, F., Greco, M., Theodoridis, E., Freedman, J., Gee, S., Chan, J.N.E., Ryan, S., Bugallo-Blanco, E., Peterson, P., Kisand, K., Haljasmägi, L., Chadli, L., Moingeon, P., Martinez, L., Merrick, B., Bisnauthsing, K., Brooks, K., Ibrahim, M.A.A., Mason, J., Lopez Gomez, F., Babalola, K., Abdul-Jawad, S., Cason, J., Mant, C., Seow, J., Graham, C., Doores, K.J., Di Rosa, F., Edgeworth, J., Shankar-Hari, M., Hayday, A.C. (2020) A dynamic COVID-19 immune signature includes associations with poor prognosis. Nat. Med. doi: 10.1038/s41591-020-1038-6. Lee, H.J., Ko, J.H., Kim, H.J., Jeong, H.J., Oh, J.Y. (2020) Mesenchymal stromal cells induce distinct myeloid-derived suppressor cells in inflammation. JCI Insight. 5(12), e136059.

Li, L., Zhang, W., Hu, Y., Tong, X., Zheng, S., Yang, J., Kong, Y., Ren, L., Wei, Q., Mei, H., Hu, C., Tao, C., Yang, R., Wang, J., Yu, Y., Guo, Y., Wu, X., Xu, Z., Zeng, L., Xiong, N., Chen, L., Wang, J., Man, N., Liu, Y., Xu, H., Deng, E., Zhang, X., Li, C., Wang, C., Su, S., Zhang, L., Wang, J., Wu, Y., Liu, Z. (2020) Effect of Convalescent Plasma Therapy on Time to Clinical Improvement in Patients With Severe and Life-threatening COVID-19: A Randomized Clinical Trial. JAMA. 324(5), 460-470.

Lucas, C., Wong, P., Klein, J., Castro, T.B.R., Silva, J., Sundaram, M., Ellingson, M.K., Mao, T., Oh, J.E., Israelow, B., Takahashi, T., Tokuyama, M., Lu, P., Venkataraman, A., Park, A., Mohanty, S., Wang, H., Wyllie, A.L., Vogels, C.B.F., Earnest, R., Lapidus, S., Ott, I.M., Moore, A.J., Muenker, M.C., Fournier, J.B., Campbell, M., Odio, C.D., Casanovas-Massana, A.; Yale IMPACT Team, Herbst, R., Shaw, A.C., Medzhitov, R., Schulz, W.L., Grubaugh, N.D., Dela Cruz, C., Farhadian, S., Ko, A.I., Omer, S.B., Iwasaki, A. (2020) Longitudinal analyses reveal immunological misfiring in severe COVID-19. Nature. 584(7821), 463–469.

Rubin, R. (2020) Testing an Old Therapy Against a New Disease: Convalescent Plasma for COVID-19. JAMA. 323, 2114–2117.

Stakenborg, M., Verstockt, B., Meroni, E., Goverse, G., Simone, V.D., Verstockt, S., Matteo, M.D., Czarnewski, P., Villablanca, E.J., Ferrante, M., Boeckxstaens, G.E., Mazzone, M., Vermeire, S., Matteoli, G.. (2020) Neutrophilic HGF-MET signaling exacerbates intestinal inflammation. J. Crohns Colitis. Jjaa121.

Tan, C.W., Chia, W.N., Qin, X., Liu, P., Chen, M.I-C., Tiu, C., Hu, Z., Chih-Wei Chen, V., Young, B.E., Sia, W.R., Tan, Y-Z., Foo, R., Yi, Y., Lye, D.C., Anderson, D.E., Wang, L-F. (2020) A SARS-CoV-2 surrogate virus neutralization test based on antibody-mediated blockage of ACE2-spike protein-protein interaction. Nat Biotechnol. 38(9), 1073–1078.

Waggoner, S.N., Reighard, S.D., Gyurova, I.E., Cranert, S.A., Mahl, S.E., Karmele, E.P., McNally, J.P., Moran, M.T., Brooks, T.R., Yaqoob, F., Rydyznski, C.E. (2016) Roles of natural killer cells in antiviral immunity. Curr. Opin. Virol. 16, 15–23.

WHO Working Group on the Clinical Characterisation and Management of COVID-19 infection. (2020) A minimal common outcome measure set for COVID-19 clinical research. Lancet Infect. Dis. 20(8), e192–e197.

